# Diagnosis of COVID-19 from X-rays Using Combined CNN-RNN Architecture with Transfer Learning

**DOI:** 10.1101/2020.08.24.20181339

**Authors:** Mabrook S. Al-Rakhami, Md. Milon Islam, Md. Zabirul Islam, Amanullah Asraf, Ali Hassan Sodhro, Weiping Ding

## Abstract

The confrontation of COVID-19 pandemic has become one of the promising challenges of the world healthcare. Accurate and fast diagnosis of COVID-19 cases is essential for correct medical treatment to control this pandemic. Compared with the reverse-transcription polymerase chain reaction (RT-PCR) method, chest radiography imaging techniques are shown to be more effective to detect coronavirus. For the limitation of available medical images, transfer learning is better suited to classify patterns in medical images. This paper presents a combined architecture of convolutional neural network (CNN) and recurrent neural network (RNN) to diagnose COVID-19 from chest X-rays. The deep transfer techniques used in this experiment are VGG19, DenseNet121, InceptionV3, and Inception-ResNetV2. CNN is used to extract complex features from samples and classified them using RNN. The VGG19-RNN architecture achieved the best performance among all the networks in terms of accuracy in our experiments. Finally, Gradient-weighted Class Activation Mapping (Grad-CAM) was used to visualize class-specific regions of images that are responsible to make decision. The system achieved promising results compared to other existing systems and might be validated in the future when more samples would be available. The experiment demonstrated a good alternative method to diagnose COVID-19 for medical staff.

## 1. Introduction

The COVID-19 outbreak has spread rapidly due to person to person transmission and created a devastating impact on global health. So far, COVID-19 has infected the world with over 86,989,000 infections and close to 1,879,176 deaths [1]. Healthcare systems have been broken down in all countries due to the limited number of intensive care units (ICUs). Coronavirus infected patients with serious conditions are admitted into ICUs. To control this pandemic, many national governments have presented ‘lockdown’ to ensure the ‘social distancing’ and ‘isolation’ among the people to reduce person to person transmission [2]. The coronavirus symptoms may vary from cold to fever, acute respiratory illness, and shortage of breath [3]. The most crucial step is to diagnose the COVID-19 at an early stage and isolated the infected people from others. RT-PCR is considered as a key indicator to diagnose COVID-19 cases reported by the government of China [4]. However, it is a time-consuming method with a high false negatives rate [5]. In many cases, the coronavirus affected may not be identified correctly for the low sensitivity.

To combat this pandemic, a lot of interest has been found in the role of medical imaging modalities [6]. The chest radiographs such as chest X-ray and computed tomography (CT) are suitable for the detection of COVID-19 due to high sensitivity that is already explored as a standard diagnosis system for pneumonia disease [7]. CT scan is more accurate than chest X-ray to diagnose pneumonia but still chest X-ray is effective due to cheaper, quicker, and fewer radiation systems [8]. Deep learning [9]-[11] is widely used in the medical field for the analysis of complex medical images. Deep learning techniques rely on automated extracted features instead of manual handcrafted features.

We proposed a combination of CNN and RNN framework to identify coronavirus cases from chest X-rays in this paper. The RNN network is capable of handling long term dependencies using internal memories. In the case of fully connected networks, nodes between layers are connectionless that process only one input but in RNN, nodes are connected from a directed graph that process an input with a specific order [12], [13], [14]. We comparatively used four very popular pre-trained CNN models namely VGG19, DenseNet121, InceptionV3, and Inception-ResNetV2 with RNN to find out the best CNN-RNN architecture within the limitations of the datasets. In this system, we first used the pre-trained CNN to extract significant features from images. Then, we applied RNN classifier to identify COVID-19 cases using the extracted features.

The contributions of this paper are summarized in the following.

i. We developed and trained a combined four CNN-RNN architectures to classify coronavirus infection from others.
ii. To detect coronavirus cases, a total of 6396 X-ray samples are used as a dataset from several sources.
iii. An exhaustive experimental analysis is ensured to measure the performance of each architecture in terms of area under the receiver operating characteristics (ROC) curve (AUC), accuracy, precision, recall, F1-score, and confusion matrix and also applied Grad-CAM to visualize the infected region of X-rays.

The paper is organized as follows. A brief review of related works is presented in Section 2. The methods and materials including dataset collection, the development of combined networks, and performance evaluation metrics are described in Section 3. Extensive result analysis with relative discussions is illustrated in Section 4. Finally, the conclusion of the paper is drawn in Section 5.

## 2. Related Works

Because of the COVID-19 pandemic, many efforts have been explored to develop a system for the diagnosis of COVID-19 using artificial intelligence techniques such as machine learning [15], deep learning [16]. In this section, a detailed description is provided about the recently developed systems to diagnose COVID-19 cases.

Luz et al. [17] introduced an extended EfficientNet model based on convolutional network architecture to analyse the lung condition using X-ray images. The model used 183 samples of COVID-19 and achieved 93.9% accuracy and 80% sensitivity for coronavirus classification. Rahimzadeh and Attar [18] presented a concatenated Xception and ResNet50V2 network to find out the infected region of COVID-19 patients from chest X-rays. The network trained on eight phases and used 633 samples in each phase including 180 samples of COVID-19. The network obtained 99.56% accuracy and 80.53% recall to detect coronavirus infection. Minaee et al. [19] illustrated a deep transfer learning architecture utilizing 71 COVID-19 samples to identify infected parts from other lung diseases. The architecture obtained an overall 97.5% sensitivity and 90% specificity to differentiate coronavirus cases. Punn and Agarwal [20] demonstrated a deep neural network to identify coronavirus symptoms. The scheme used 108 COVID-19 cases and obtained an average 97% accuracy. Khan et al. [21] introduced a deep CNN to diagnose coronavirus disease from 284 COVID-19 samples. The proposed framework found an accuracy of 89.5%, and a precision of 97% to detect coronavirus. Wang and Wong [22] presented COVID-Net to distinguish COVID-19 cases from others using chest X-ray samples. The system achieved 92.4% accuracy by analysing 76 samples of COVID-19. Narin et al. [23] proposed a deep transfer learning with three CNN architectures and used a small dataset including 50 chest X-rays for both COVID-19 and normal cases to detect coronavirus infection. The ResNet50 showed high performance with 98.6% accuracy, 96% recall, and 100% specificity among other networks.

Hemdan et al. [24] developed COVIDX-Net framework including seven pre-trained CNN to detect coronavirus infection from X-ray samples. The dataset consisted of 25 samples of COVID-19 cases and 25 samples of normal cases. The framework obtained high performance for VGG19 with 0.89 F1-score. Apostolopoulos and Mpesiana [25] presented a transfer learning scheme for the detection of coronavirus infection. The VGG19 obtained high performance among others with an accuracy of 93.48%, specificity of 92.85%, and sensitivity of 98.75%. Horry et al. [26] illustrated deep transfer learning based system and achieved the highest result for VGG19 with 83% recall and 83% precision for the diagnosis of COVID-19. Loey et al. [27] proposed a deep transfer learning approach with three pre-trained CNN networks to diagnose coronavirus disease. The dataset includes 69 COVID-19 samples, 79 pneumonia bacterial samples, 79 pneumonia virus samples, and 79 normal samples. The GoogleNet achieved an accuracy of 80.6% in the four cases scenario. Kumar and Kumari [28] used a transfer learning based system using nine pre-trained CNNs combined with support vector machine (SVM) to classify coronavirus infected patients. The ResNet50-SVM is showed the best performance among other models with an accuracy of 95.38%. Bukhari et al. [29] proposed a transfer learning technique for the detection of COVID-19 from X-ray samples. The system used 89 samples of COVID-19 and obtained 98.18% accuracy with 98.19% F1-score. Abbas et al. [30] introduced a DeTraC architecture to detect coronavirus from 105 samples of COVID-19. The architecture achieved 95.12% accuracy, 91% sensitivity, 91.87% specificity, and 93.36% precision to diagnose coronavirus infection. Islam et al. [31] applied a combined CNN and LSTM architecture to classify coronavirus cases using X-ray images. The scheme applied 421 samples including 141 COVID-19 cases and achieved an accuracy of 97%, specificity of 91%, and sensitivity of 93%.

## 3. Methods and Materials

Though some of the existing systems showed promising results, COVID-19 dataset was quite small, and also the variable quality of these datasets was not addressed. It also noticed that the used dataset in those experiments was quite unbalanced that could lead to over-classification of majority class at the expense of under-classification of the minority class. On the contrary, COVID-19 images were highly inconsistent as they were retrieved from the different regions of the world whereas pneumonia and normal images were uniform as well as highly curated in previous studies. Here, COVID-19 dataset contained most adult’s patients, and the pneumonia dataset used mostly young patients. These discrepancies were mostly ignored in the existing systems. Therefore, our proposed system used a balanced dataset with adults and young patient’s images to learn the actual features of the disease. The proposed system contains several steps to diagnose COVID-19 infection that is shown in Fig. 1. Firstly, in the preprocessing pipeline, chest X-ray samples were resized, shuffled, and normalized to figure out the actual features and reduce the unwanted noise from the images. Afterward, the dataset was partitioned into training and testing sets. Using the training dataset, we applied four pre-trained CNN architecture combined with RNN classifier. The accuracy and loss of training datasets were obtained after each epoch and using five-fold cross-validation technique, the validation loss and accuracy were found. The performance of the overall system was measured with a confusion matrix, accuracy, precision, recall, AUC, and F1-score metrics.

**Fig 1.**
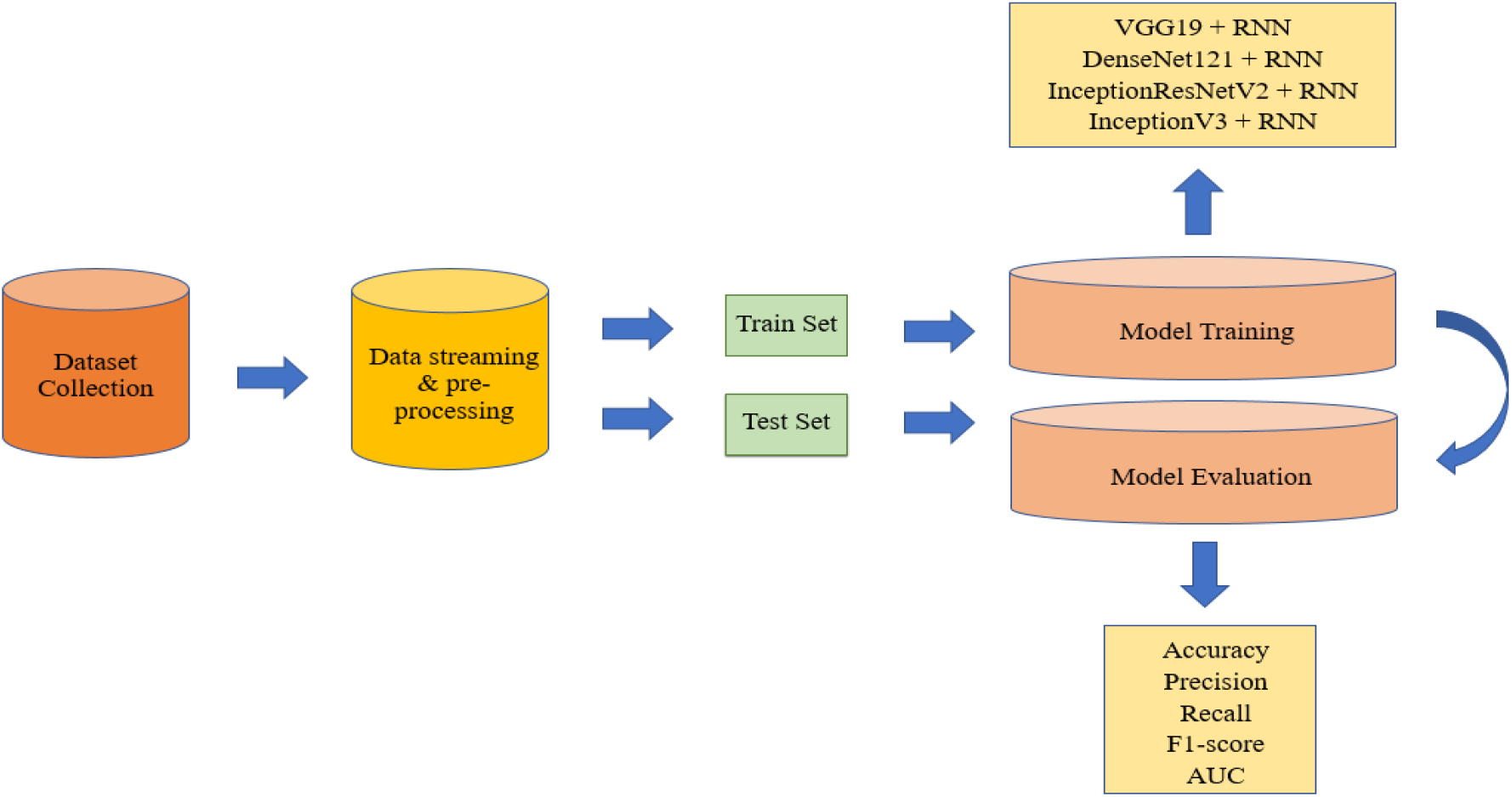
The overall system architecture of the COVID-19 diagnosis framework

### 3.1. Experimental Dataset

In this paper, X-ray samples of COVID-19 were retrieved from seven different sources for the unavailability of a large specific dataset. Firstly, a total 1401 samples of COVID-19 were collected using GitHub repository [32], [33], the Radiopaedia [34], Italian Society of Radiology (SIRM) [35], Figshare data repository websites [36], [37]. Then, 912 augmented images were also collected from Mendeley instead of using data augmentation techniques explicitly [38]. Finally, 2313 samples of normal and pneumonia cases were obtained from Kaggle [39], [40]. A total of 6939 samples were used in the experiment, where 2313 samples were used for each case. The total dataset was divided into 80%-20% for training and testing sets where 1850 samples of COVID-19, 1851 samples of pneumonia, and 1850 samples of normal cases were used for training including all augmented images. The remaining 463 samples of COVID-19, 462 samples of pneumonia, and 463 samples of normal cases were used for the testing including only original images; no augmented images were used here. Pixel normalization was applied to images in data preprocessing step.

### 3.2. Development of Combined Network

#### 3.2.1. Deep Transfer Learning with CNN

Transfer Learning [41] is an approach where information extracted by one domain is transferred to another related domain. It is applied when the dataset is not sufficient to train the parameters of any network. In this part, four pre-trained CNNs are described to accomplish the proposed CNN-RNN architecture as follows. In addition, the characteristics of four pre-trained CNN architectures are shown in Table 1.

**Table 1.** Characteristics of four pre-trained CNN architectures

| Network | Depth | Parameters ($10^6$) |
| --- | --- | --- |
| VGG19 | 19 | 144.0 |
| DenseNet121 | 121 | 8.0 |
| InceptionV3 | 48 | 23.6 |
| Inception-ResNetV2 | 164 | 56.0 |

i. VGG19: VGG19 [42] is a version of the visual geometry group network (VGG) developed by Karen Simonyan and Andrew Zisserman based on deep network architecture. It has 19 layers in total including 16 convolutional layers with three fully-connected layers to perform on the ImageNet dataset [43]. VGG19 used a 3 × 3 convolutional filter and a stride of 1 that was followed by multiple non-linear layers. Max-pooling is applied in VGG19 to reduce the volume size of the image and achieved high accuracy on image classification.
ii. DenseNet121: Dense Convolutional Network (DenseNet) [44] uses dense connections instead of direct connections among the hidden layers developed by Huang et al. In DenseNet architecture, each layer is connected to the next layer to transfer the information among the network. The feature maps are transmitted directly to all subsequent layers and use only a few parameters for training. The overfitting of a model is reduced by dense connections for small datasets. DenseNet121 has 121 layers, loaded with weights from the ImageNet dataset.
iii. InceptionV3: InceptionV3 [45] is used to improve computing resources by increasing the depth and width of the network. It has 48 layers with skipped connections to use a building block and trained on million images including 1000 categories. The inception module is repeated with max-pooling to reduce dimensionality.
iv. Inception-ResNetV2: Inception-ResnetV2 [46] network is a combination of inception structure with residual connections including 164 deep layers. It has multiple sized convolution filters trained on millions of images and avoids the degradation problem.

#### 3.2.2. Recurrent Neural Network

Recurrent neural network [47] is an extended feedforward neural network with one or more feedback loops designed for processing sequential data. Given, an input sequence (x_1_, …, x_t_), a RNN generates output sequence of (y_1_, …, y_t_) by using the following formulas and RNN structure is shown in Fig. 2.

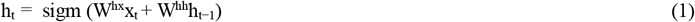

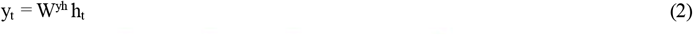

**Fig. 2.**
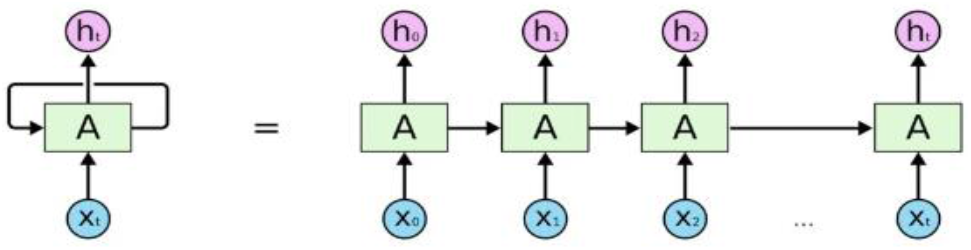
The structure of recurrent neural networks

RNN is used whenever the input-output relationship is found based on time and capacity to handle long term dependencies [48]. The strategy of modeling sequence is to feed the input sequence to a fixed-sized vector using a RNN, and then to map the vector to a softmax layer. Unfortunately, a problem occurs in RNN when the gradient vector is increasing and decreasing exponentially for long sequences. This vanishing gradient and exploding problem [49] create difficulties to learn long-range relationships from the sequences of the RNN architecture. However, the Long Short-Term Memory (LSTM) [50] is capable to solve such a long-distance dependencies problem successfully. The main difference from RNN is that LSTM added a separate memory cell state to store long term states and updates or exposes them whenever necessary. The LSTM consists of three gates: input gate, forget gate, and output gate where *i*_*t*_ denotes input gate, f_*t*_ denotes forget gate, O_*t*_ denotes output gate, 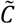 cell input activation vector, c_t_ refers to the memory cell state, and *h*_*t*_ refers to the hidden state at each time step t.

The transition representations of LSTM are as follows.

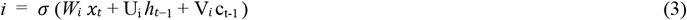

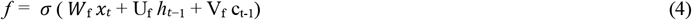

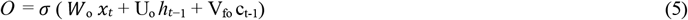

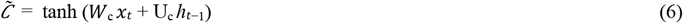

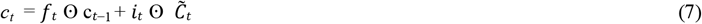

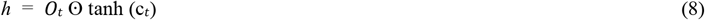

where x_t_ refers current input, *σ* refers sigmoid function and ⊙ refers element wise multiplication.

#### 3.2.3. Development of CNN-RNN Hybrid Network

A combined method using CNN and RNN was developed for the diagnosis of COVID-19 using three types of X-ray samples in this paper. The complex features were extracted from 224 × 224 × 3 sized samples using VGG19, DeneNet121, InceptionV3, and Inception-ResNetV3. The extracted features were feed to the single layered RNN classifier i.e. the output is produced by passing it through a single hidden state to differentiate COVID-19, pneumonia, and normal cases. The dimensionality of feature maps of pretrained CNN and how CNN is connected to RNN were shown in Table 2.

**Table 2.** The summary of the used architectures. (a) VGG19-RNN (b) DenseNet121-RNN (c) Inception-ResNetV2-RNN (d) InceptionV3-RNN

| Layer (type) | Input Size | Output Size |
| --- | --- | --- |
| pretrained (VGG19) | 224, 224, 3 | 7, 7, 512 |
| reshape (Reshape) | 7, 7, 512 | 49, 512 |
| lstm (LSTM) | 49, 512 | 49, 512 |
| flatten (Flatten) | 49, 512 | 25088 |
| fc_1 (Dense) | 25088 | 4096 |
| fc_2 (Dense) | 4096 | 4096 |
| output (Dense) | 4096 | 3 |
| (a) |  |  |
| Layer (type) | Input Size | Output Size |
| pretrained (DenseNet121) | 224, 224, 3 | 7, 7, 1024 |
| reshape (Reshape) | 7, 7, 1024 | 49, 1024 |
| lstm (LSTM) | 49, 1024 | 49, 1024 |
| flatten (Flatten) | 49, 1024 | 50176 |
| fc_1 (Dense) | 50176 | 4096 |
| fc_2 (Dense) | 4096 | 4096 |
| output (Dense) | 4096 | 3 |
| (b) |  |  |
| Layer (type) | Input Size | Output Size |
| pretrained (Inception-ResNetV2) | 224, 224, 3 | 5, 5, 1536 |
| reshape (Reshape) | 5, 5, 1536 | 25, 1536 |
| lstm (LSTM) | 25, 1536 | 25, 512 |
| flatten (Flatten) | 25, 512 | 12800 |

| fc_1 (Dense) | 12800 | 4096 |
| --- | --- | --- |
| fc_2 (Dense) | 4096 | 4096 |
| output (Dense) | 4096 | 3 |
| (c) |  |  |
| Layer (type) | Input Size | Output Size |
| pretrained (InceptionV3) | 224, 224, 3 | 5, 5, 2048 |
| reshape (Reshape) | 5, 5, 2048 | 25, 2048 |
| lstm (LSTM) | 25, 2048 | 25, 512 |
| flatten (Flatten) | 25, 512 | 12800 |
| fc_1 (Dense) | 12800 | 4096 |
| fc_2 (Dense) | 4096 | 4096 |
| output (Dense) | 4096 | 3 |
| (d) |  |  |

The CNN-RNN network for COVID-19 classification is shown in Fig. 3 which contains the following steps.

**Fig. 3.**
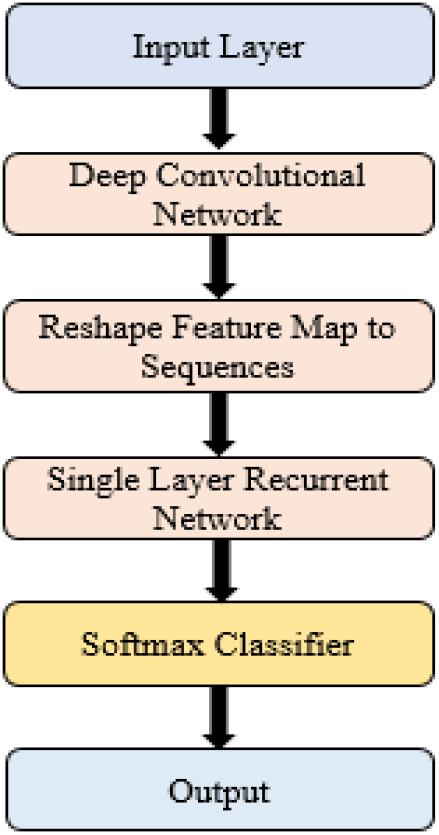
The workflow of the CNN-RNN architecture for COVID-19 diagnosis

Step 1: Use different pre-trained CNN models to extract essential features from X-ray images.

Step 2: Reshape the feature map into the sequence.

Step 3: Set the feature map as the input of single-layered RNN.

Step 4: Apply a softmax classifier to classify COVID-19 X-ray images.

In transfer learning, the activations of convolutional layers are the same as original architectures. Finally, the fully connected layers were activated using Rectified Linear Unit (ReLU) [51] and Dropout layer [52] was used in RNN layers to prevent overfitting [53] of the models. All the layers of pretrained CNN were frozen during training except RNN and fully connected layers. Finally, the CNN-RNN architectures were trained with RMSprop [54] and the batch size of 32, the learning rate of 0.00001, and a total of 100 epochs were conducted for training. The samples were shuffled in batches between epochs. We used single layer RNN combined with pretrained CNN shown in Fig. 3. In a single layer RNN, the output is produced by passing it through a single hidden state to capture structure of a sequence. We have used RNN as a sequence-to-sequence layer and take output sequence as an input for fully-connected layers downstream in the developed system. In this architecture, Gradient clipping is used to handle the long sequence problem. In our proposed system, the order of elements of a sequence is horizontal. Basically, to develop the sequence by collecting the pixels from the images in three orders such as horizontal, vertical and spiral are used. The structure of the combined CNN-RNN is shown in Fig. 4. Algorithm 1 presented the proposed CNN-RNN technique to detect COVID-19 cases.

**Fig. 4.**
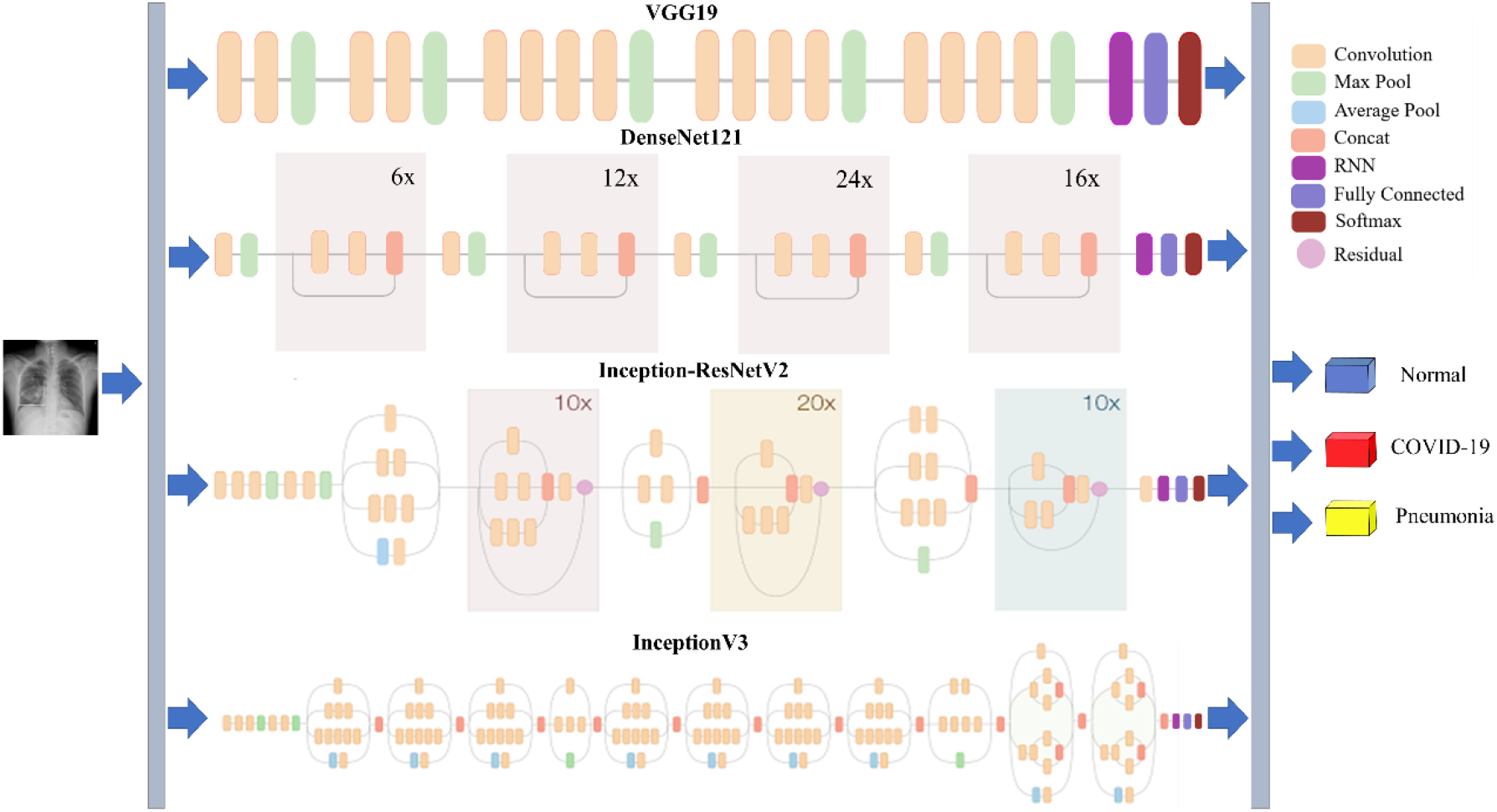
The structure of the combined CNN-RNN architecture for COVID-19 diagnosis

##### Algorithm 1

CNN-RNN Algorithm

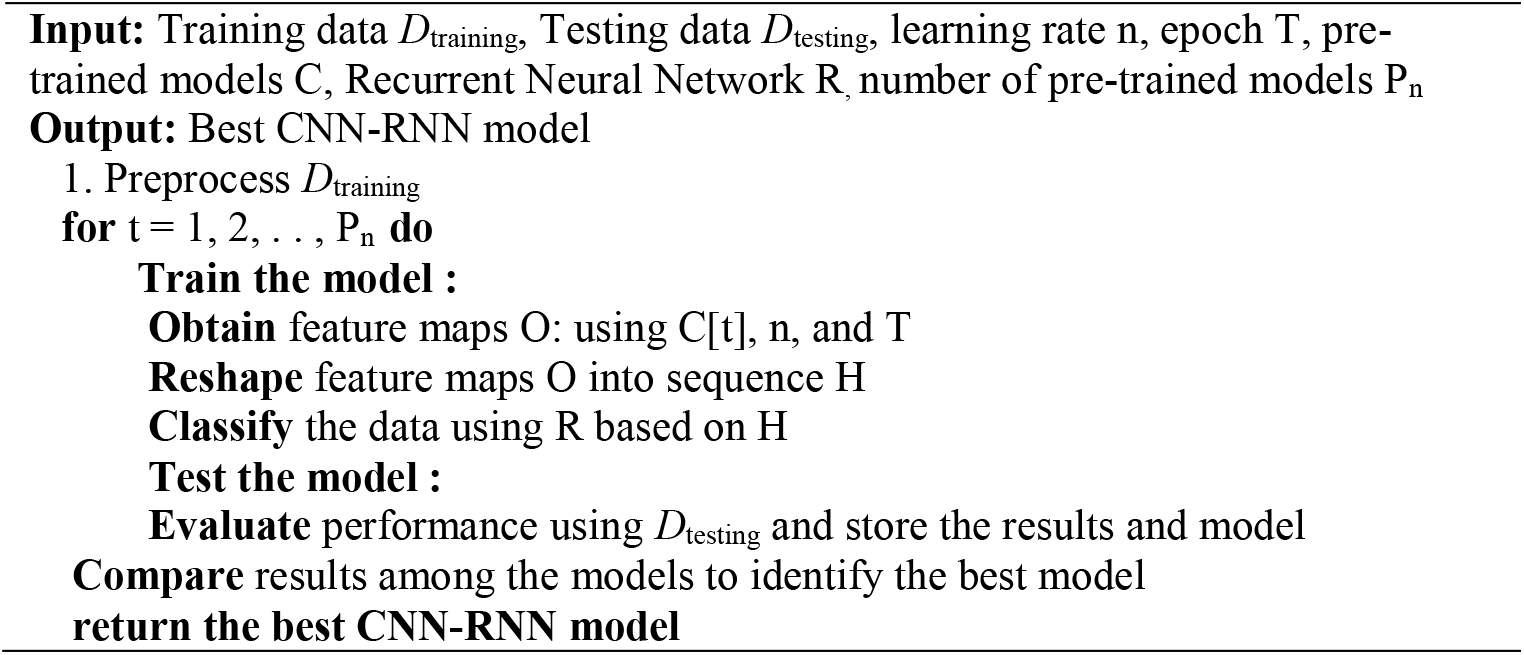

### 3.3 Evaluation Criteria

The performance of the developed system is measured in terms of AUC, accuracy, precision, recall, and F1-score. The evaluation metric parameters are represented mathematically in the following. Here, correctly classified COVID-19 cases are denoted by True Positive (TP), correctly classified pneumonia or normal cases are represented by True Negative (TN), wrongly classified as COVID-19 cases are denoted by False Positive (FP) and wrongly classified as pneumonia or normal cases are depicted by False Negative (FN).

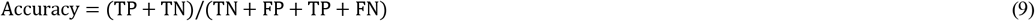

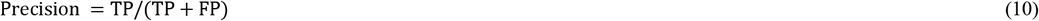

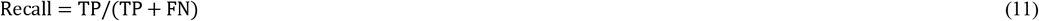

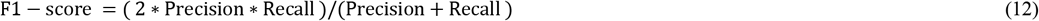

All the experiments were performed on a Google Colaboratory Linux server with Ubuntu 16.04 operating system using Tesla K80 GPU graphics card and the TensorFlow/Keras framework of python language.

## 4. Results Analysis

### 4.1. Results Analysis

The accuracy and loss curves in the training and validation phases are shown in Fig. 5. For VGG19-RNN architecture, the highest training and validation accuracy is observed 99.01% and 97.74% and loss is 0.02 and 0.09 at epoch 100. On the contrary, the lowest training and validation accuracy is obtained 98.03% and 94.91% and loss is 0.05 and 0.26 at epoch 100 for the InceptionV3-RNN network. Analyzing the loss curve, it is seen that the loss values of VGG19-RNN decrease faster and tends to zero than other networks.

**Fig. 5.**
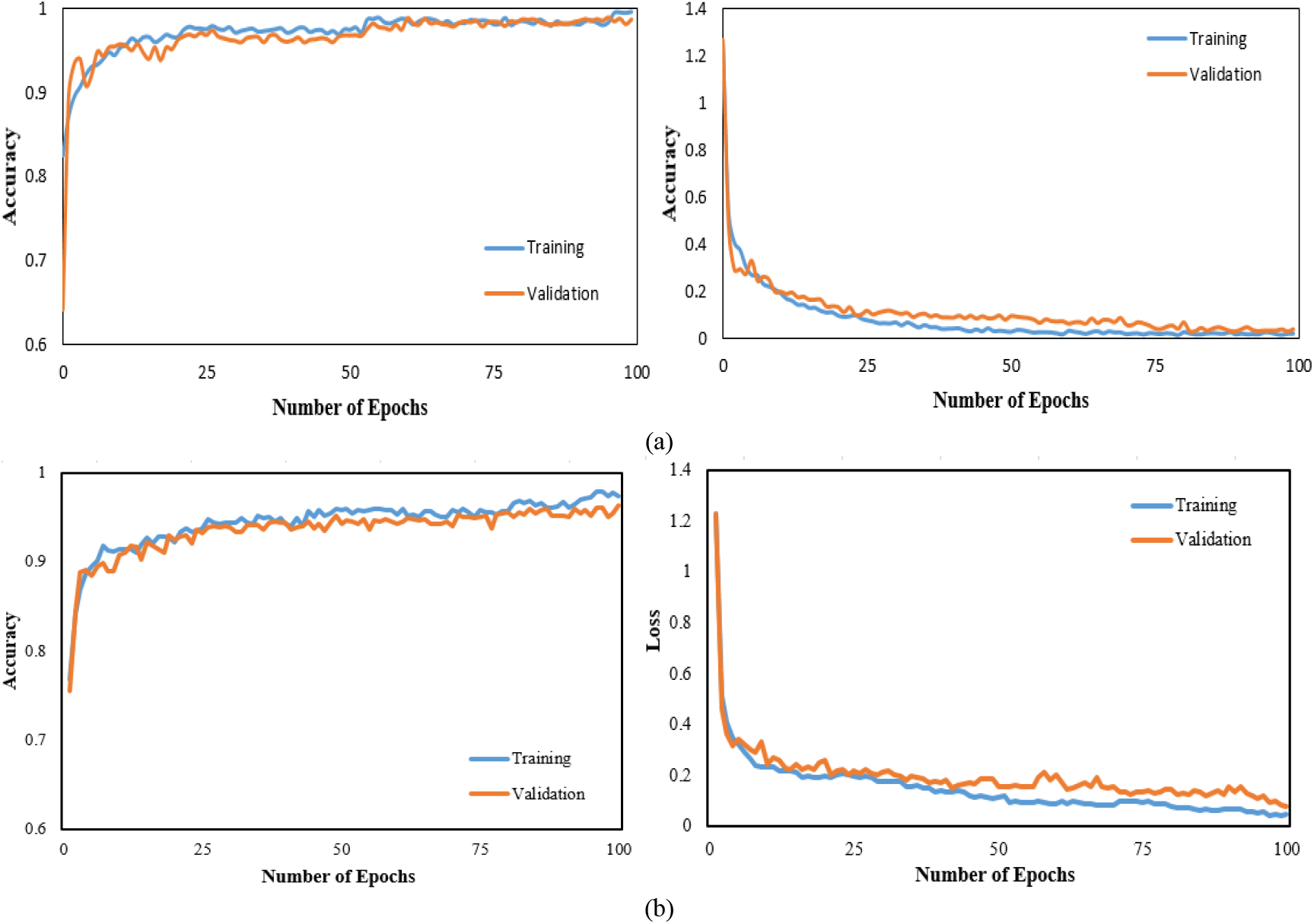
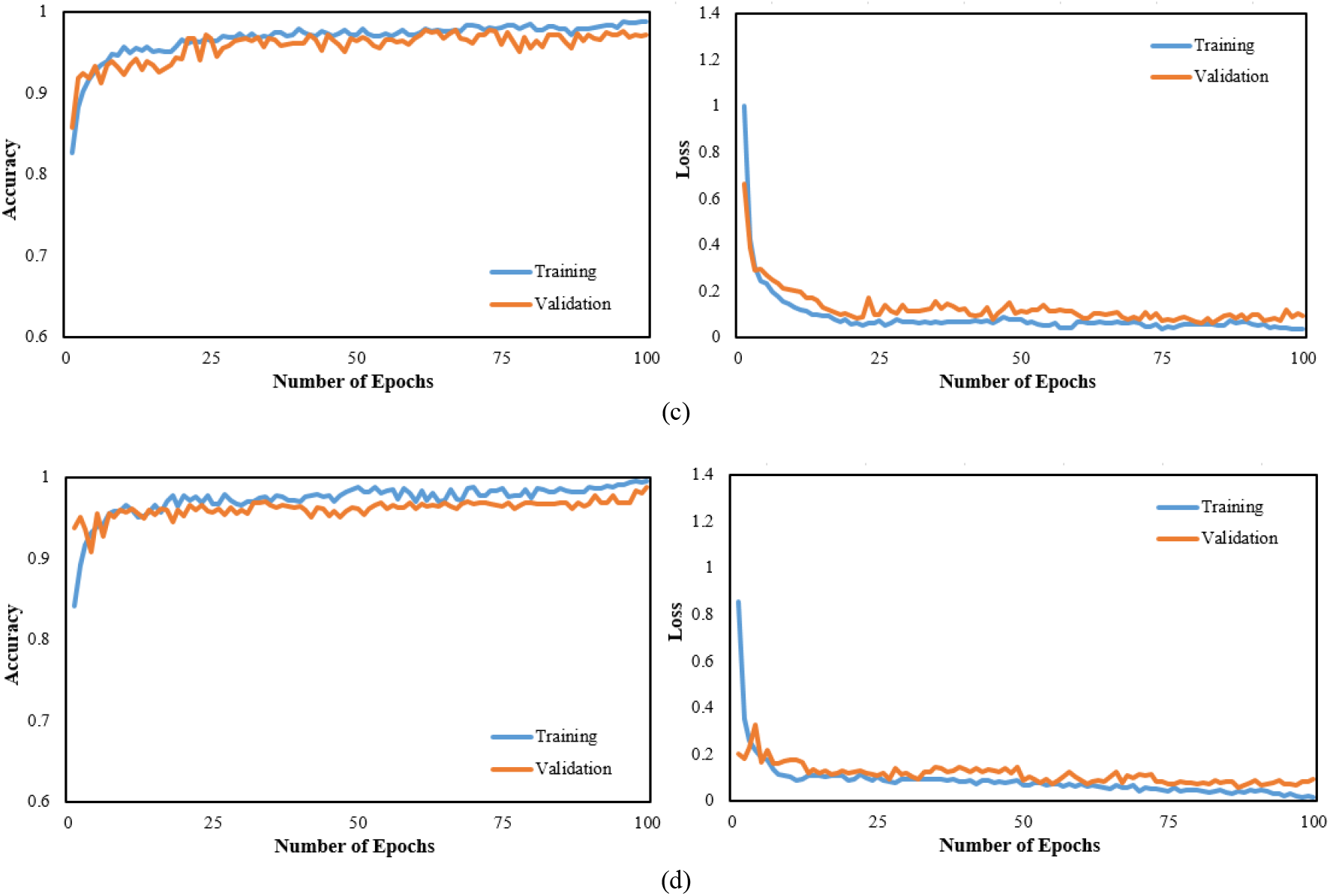
Accuracy and loss curve of four CNN-RNN architectures. (a) VGG19 (b) DenseNet121(c) InceptionV3 (d) Inception-ResNetV2

Fig. 6 demonstrates the confusion matrix of the developed architectures. Among 1388 samples, 2 samples were misclassified by VGG19-RNN network including only one sample for COVID-19 cases, 3 samples were misclassified by DenseNet121-RNN network including two COVID-19 samples, 20 samples were misclassified by InceptionV3-RNN architecture consisting of three COVID-19 samples and 7 samples were misclassified by the Inception-ResNetV2-RNN network comprising of seven COVID-19 samples. Hence, it was found that VGG19-RNN architecture is superior to other networks and selected as a main deep learning architecture with high performance.

**Fig. 6.**
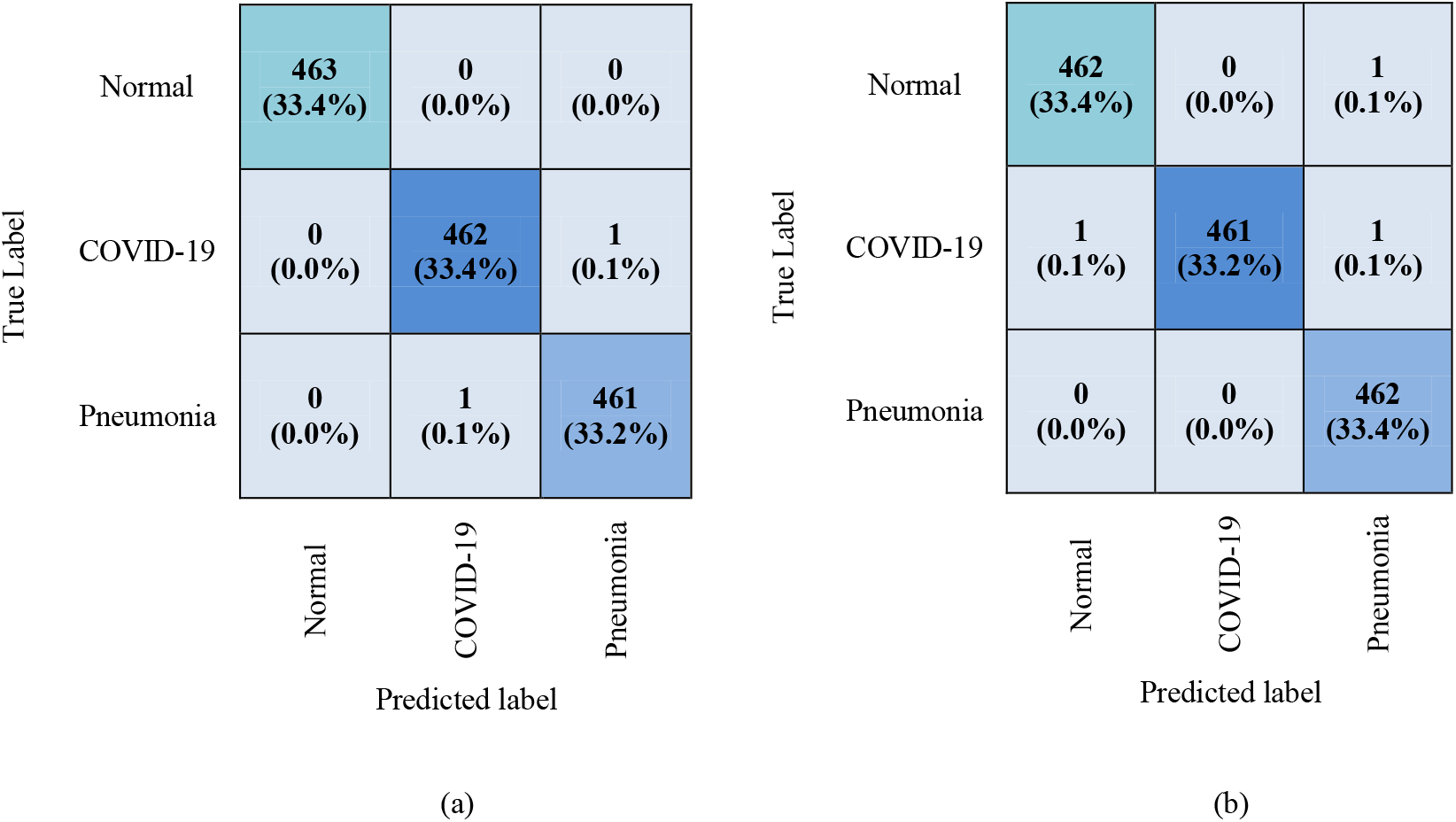
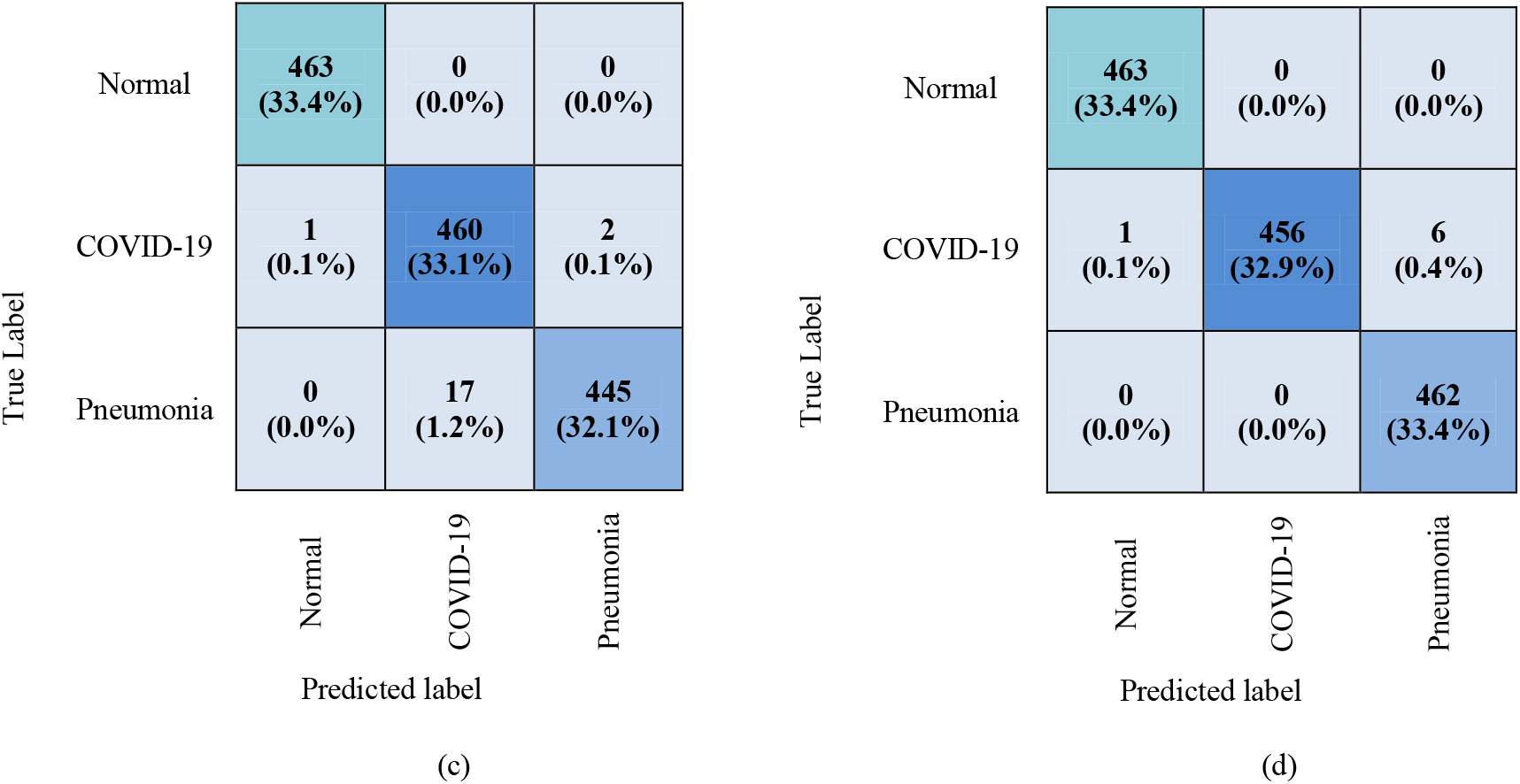
Confusion matrix of the CNN-RNN architecture for COVID-19 diagnosis. (a) VGG19 (b) DenseNet121 (c) InceptionV3 (d) Inception-ResNetV2

In this paper, the performance of four CNN-RNN architectures is summarized in Table 3. The best performance was found by the VGG19-RNN network with 99.86% accuracy, 99.99% AUC, 99.78% precision, 99.78% recall, and 99.78% F1-score for COVID-19 cases. On the contrary, the comparatively low performance was obtained by InceptionV3-RNN architecture with 98.56% accuracy, 99.95% AUC, 99.35% precision, 96.44% recall, and 97.87% F1-score. Besides, ROC curves also added between TP and FP rate for all networks shown in Fig. 7. The networks can differentiate COVID-19 cases from others with AUC in the range of 99.95% to 99.99%. For better visualization and to show the differences between the classifiers Precision-Recall (PR) curve is also added shown in Fig. 8.

**Table 3.** Performance of the combined CNN-RNN architecture

| Classifier | Patient Status | AUC (%) | Accuracy (%) | Precision (%) | Recall (%) | F1-score (%) |
| --- | --- | --- | --- | --- | --- | --- |
| VGG19-RNN | COVID-19 | 99.99 | 99.86 | 99.78 | 99.78 | 99.78 |
|  | Pneumonia |  | 99.86 | 99.78 | 99.78 | 99.78 |
|  | Normal |  | 99.86 | 100.0 | 100.0 | 100.0 |
| DenseNet121-RNN | COVID-19 | 99.99 | 99.78 | 99.57 | 100.0 | 99.78 |
|  | Pneumonia |  | 99.78 | 100.0 | 99.57 | 99.78 |
|  | Normal |  | 99.78 | 99.78 | 99.78 | 99.78 |
| InceptionV3-RNN | COVID-19 | 99.95 | 98.56 | 99.35 | 96.44 | 97.87 |
|  | Pneumonia |  | 98.56 | 96.32 | 99.55 | 97.91 |
|  | Normal |  | 98.56 | 100.0 | 99.78 | 99.89 |
| Inception-ResNetV2-RNN | COVID-19 | 99.99 | 99.50 | 98.49 | 100.0 | 99.24 |
|  | Pneumonia |  | 99.50 | 100.0 | 99.72 | 99.86 |
|  | Normal |  | 99.50 | 100.0 | 99.78 | 99.89 |

**Fig. 7.**
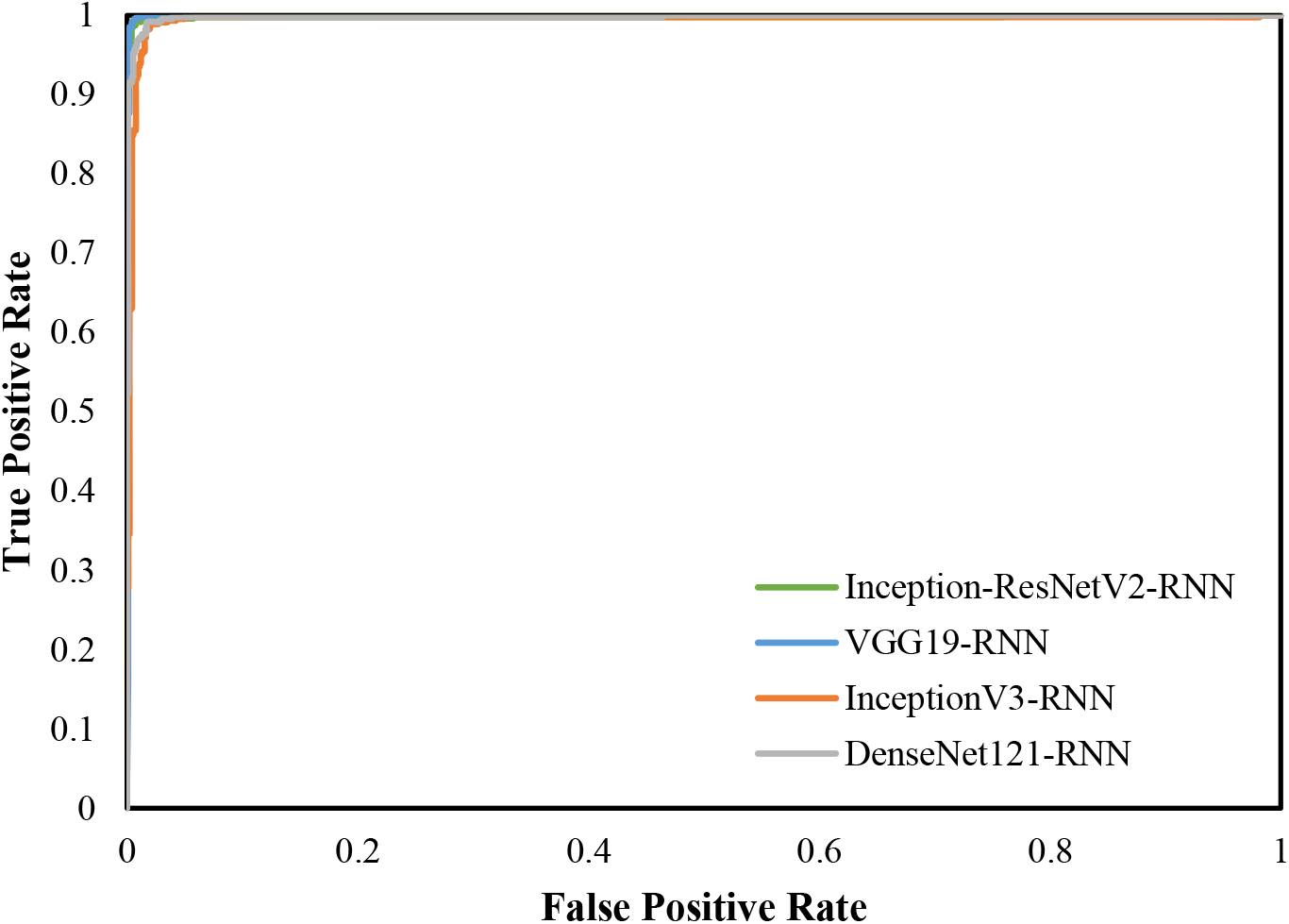
ROC curve of four combined CNN-RNN networks

**Fig. 8.**
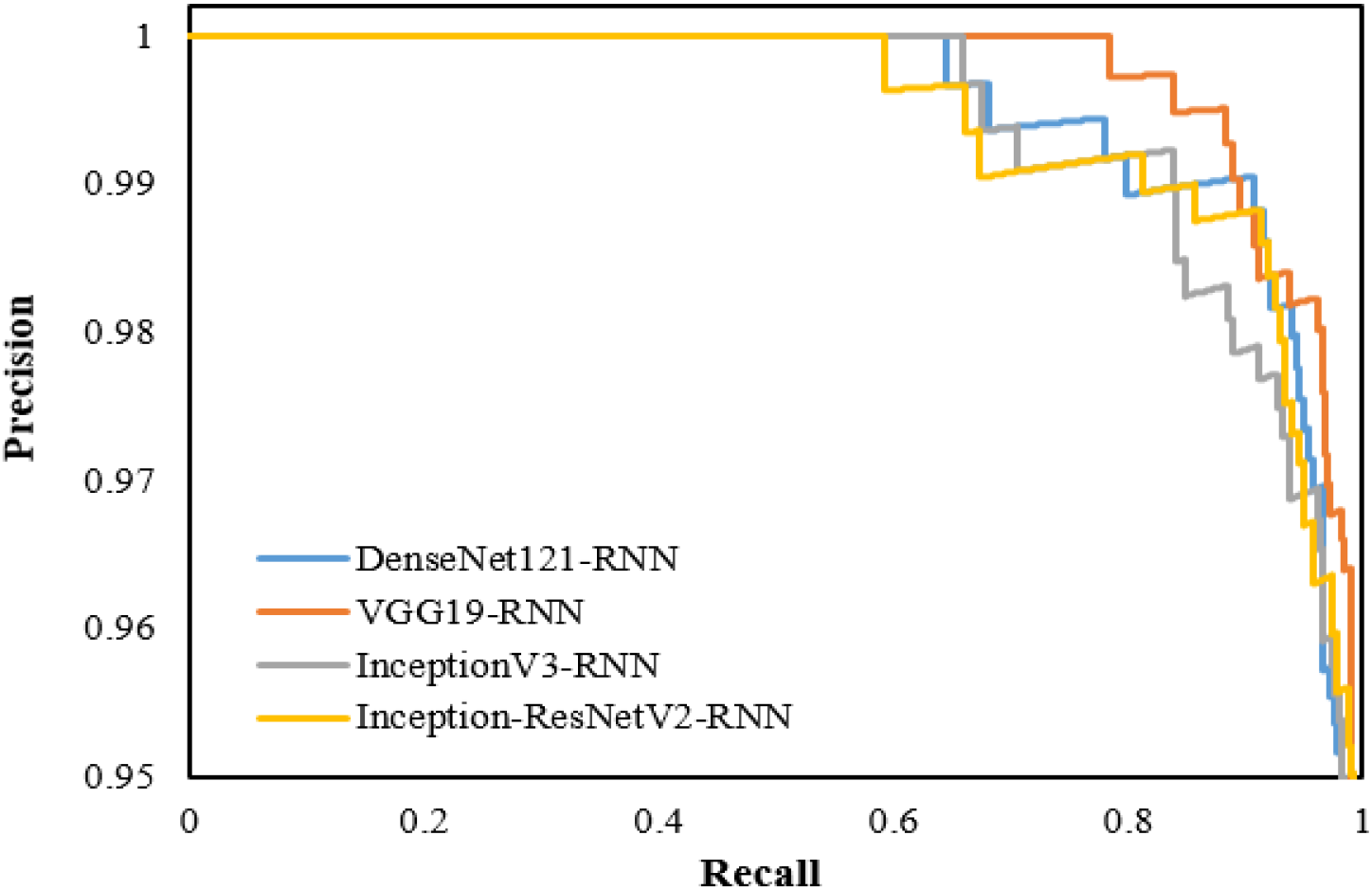
PR curve of four combined CNN-RNN networks

Finally, Grad-CAM is applied that refers to a heat-map to highlight class-specific regions of chest X-rays. Fig. 9 shows the heatmaps and superimposed images of COVID-19, pneumonia, and normal cases for the VGG19-RNN network.

**Fig. 9.**
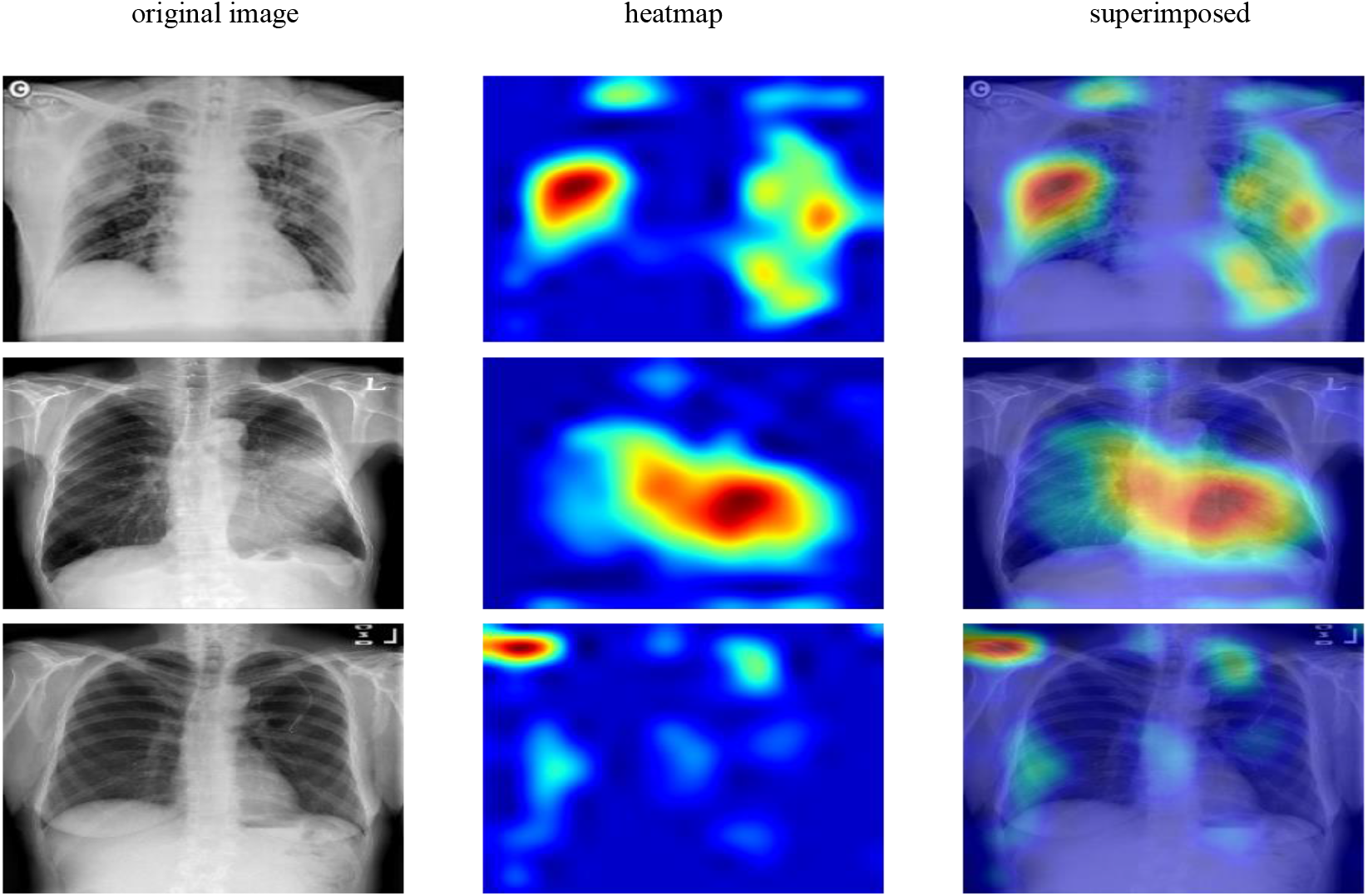
First, second and third rows represent the samples of COVID-19, pneumonia, and normal correspondingly. Besides, first, second and third columns refer to the original, heatmap and superimposed images for VGG19-RNN

### 4.2. Discussions

In this paper, the combination of four CNN and RNN was used to diagnose COVID-19 infection. The results demonstrated that VGG19-RNN is more effective to differentiate COVID-19 cases from pneumonia and normal cases and is considered as a main deep learning architecture. A comparison between simple CNN based pretrained networks with our study is demonstrated in Table 4. It is clearly shown that VGG19-RNN network has obtained higher performance than pretrained CNN networks. Finally, another comparison between recent works with our study is demonstrated in Table 5. It is observed that existing systems can distinguish coronavirus infection with accuracy in the range of 80.6% to 98.3%. On the contrary, the VGG19-RNN network obtained 99.9% accuracy which is higher than other existing systems. Hence, finally, it is evident that the VGG19-RNN network showed good performance compared to other studies.

**Table 4.** Comparison between pretrained CNN with CNN-RNN architecture based on COVID-19 patients

| Classifier | Accuracy (%) | Precision (%) | Recall (%) | F1-score (%) |
| --- | --- | --- | --- | --- |
| VGG19 | 99.63 | 99.51 | 99.65 | 99.58 |
| DenseNet121 | 99.26 | 99.45 | 99.38 | 99.41 |
| InceptionV3 | 99.13 | 98.71 | 98.92 | 98.81 |
| Inception-ResNetV2 | 99.28 | 99.38 | 98.06 | 98.72 |
| VGG19-RNN | 99.86 | 99.78 | 99.78 | 99.78 |

**Table 5.** Comparative study of the proposed CNN-RNN architecture with existing works with respect to accuracy

| Author | Architecture | Accuracy (%) | COVID-19 accuracy (%) |
| --- | --- | --- | --- |
| Luz et al. [17] | EfficientNet | 93.9 | - |
| Rahimzadeh and Attar [18] | Xception - ResNet50V2 | 91.4 | 99.6 |
| Punn and Agarwal [20] | NASNetLarge | 97.0 | - |
| Khan et al. [21] | CoroNet (Xception) | 89.5 | 96.6 |
| Wang and Wong [22] | Tailored CNN | 92.3 | 80.0 |
| Narin et al. [23] | ResNet50 | 98.6 | - |
| Hemdan et al. [24] | VGG19 | 90.0 | - |
| Apostolopoulos and Mpesiana [25] | VGG19 | 93.5 | - |
| Loey et al. [27] | GoogleNet | 80.6 | 100.0 |
| Kumar and Kumari [28] | ResNet50-SVM | 95.4 | - |
| Bukhari et al. [29] | ResNet50 | 98.2 | - |
| Abbas et al. [30] | DeTrac | 95.1 | - |
| Islam et al. [31] | CNN-LSTM | 97.0 | - |
| Ucar and Korkmaz [55] | COVIDiagnosis-Net | 98.3 | 100.0 |
| Asnaoui et al. [56] | Inception-ResNetV2 | 92.2 | - |
| Li et al. [54] | DenseNet | 88.9 | 79.2 |
| Chowdhury et al. [55] | Sgdm-SqueezeNet | 98.3 | 96.7 |
| **Proposed System** | **VGG19-RNN** | **99.9** | **99.9** |

## 5. Conclusion

During the COVID-19 pandemic, the use of deep learning techniques for the diagnosis of COVID-19 has become a crucial issue to overcome the limitation of medical resources. In this study, we introduced a combination of CNN with deep transfer learning and RNN for classifying the X-ray samples into three categories of COVID-19, pneumonia, and normal. The four popular CNN networks were applied to extract features and RNN network used to classify different classes using these features. The VGG19-RNN is considered as the best network with 99.9% accuracy, 99.9% AUC, 99.8% recall, and 99.8% F1-score to detect COVID-19 cases. Hopefully, it would reduce the workload for the doctor to test COVID-19 cases.

There are some limitations of our proposed system. First, the COVID-19 samples are small that need to be updated with more samples to validate our proposed system. Second, this experiment only works with a posterior-anterior view of chest X-ray, hence it is not able to effectively classify other views such as apical, lordotic, etc. Third, the performance of our experiment is not compared with radiologists that would be our future work.

## Data Availability

The data will be available on request.

## Data Availability Statement

The data presented in this study are available upon request to the authors.

## Acknowledgement

The authors are grateful to the Deanship of Scientific Research, King Saud University for funding through Vice Deanship of Scientific Research Chairs.

## References

[1] About Worldometer COVID-19 data - Worldometer. (Accessed 06 January, 2021).

[2] Advice for the public. https://www.who.int/emergencies/diseases/novel-coronavirus-2019/advice-for-public (Accessed July 14, 2020).

[3] Everything about the Corona virus - Medicine and Health. https://medicine-and-mentalhealth.xyz/archieves/4510. (Accessed 06 June, 2020).

[4] T. Ai, Z. Yang, L. Xia, Correlation of Chest CT and RT-PCR Testing in Coronavirus Disease, Radiology. 2019 (2020) 1–8. https://doi.org/10.14358/PERS.80.2.000.

[5] C. Long, H. Xu, Q. Shen, X. Zhang, B. Fan, C. Wang, B. Zeng, Z. Li, X. Li, H. Li, Diagnosis of the Coronavirus disease (COVID-19): rRT-PCR or CT?, Eur. J. Radiol. 126 (2020) 108961. https://doi.org/10.1016/j.ejrad.2020.108961.

[6] H. Shi, X. Han, N. Jiang, Y. Cao, O. Alwalid, J. Gu, Y. Fan, C. Zheng, Articles Radiological findings from 81 patients with COVID-19 pneumonia in Wuhan, China : a descriptive study, Lancet Infect. Dis. 20 (2020) 425–434. https://doi.org/10.1016/S1473-3099(20)30086-4.

[7] Z.Y. Zu, M. Di Jiang, P.P. Xu, W. Chen, Q.Q. Ni, G.M. Lu, L.J. Zhang, Coronavirus Disease 2019 (COVID-19): A Perspective from China, Radiology. (2020) 200490. https://doi.org/10.1148/radiol.2020200490.

[8] G.D. Rubin, C.J. Ryerson, L.B. Haramati, N. Sverzellati, J.P. Kanne, S. Raoof, N.W. Schluger, A. Volpi, J.-J. Yim, I.B.K. Martin, D.J. Anderson, C. Kong, T. Altes, A. Bush, S.R. Desai, J. Goldin, J.M. Goo, M. Humbert, Y. Inoue, H.-U. Kauczor, F. Luo, P.J. Mazzone, M. Prokop, M. Remy-Jardin, L. Richeldi, C.M. Schaefer-Prokop, N. Tomiyama, A.U. Wells, A.N. Leung, The Role of Chest Imaging in Patient Management During the COVID-19 Pandemic, Chest. (2020) 1–11. https://doi.org/10.1016/j.chest.2020.04.003.

[9] Islam MM, Karray F, Alhajj R, Zeng J. A review on deep learning techniques for the diagnosis of novel coronavirus (COVID-19). IEEE Access 9 (9): 30551–30572. https://doi.org/10.1109/ACCESS.2021.3058537

[10] Asraf, A., Islam, M.Z., Haque, M.R. et al.. Deep Learning Applications to Combat Novel Coronavirus (COVID-19) Pandemic. SN COMPUT. SCI. 1, 363 (2020). https://doi.org/10.1007/s42979-020-00383-w.

[11] P. Saha, M.S. Sadi, M. M. Islam, EMCNet: Automated COVID-19 diagnosis from X-ray images using convolutional neural network and ensemble of machine learning classifiers, Inform Med Unlocked. 22 (2020) 100505. https://doi.org/10.1016/j.imu.2020.100505

[12] J. Wang, Y. Yang, J. Mao, Z. Huang, C. Huang and W. Xu, “CNN-RNN: A Unified Framework for Multi-label Image Classification,” 2016 IEEE Conference on Computer Vision and Pattern Recognition (CVPR), Las Vegas, NV, 2016, pp. 2285–2294, doi: 10.1109/CVPR.2016.251.

[13] Q. Yin, R. Zhang, X. Shao, CNN and RNN mixed model for image classification, MATEC Web Conf. 277 (2019) 02001. https://doi.org/10.1051/matecconf/201927702001.

[14] Y. Guo, Y. Liu, E.M. Bakker, Y. Guo, M.S. Lew, CNN-RNN: a large-scale hierarchical image classification framework, Multimed. Tools Appl. 77 (2018) 10251–10271. https://doi.org/10.1007/s11042-017-5443-x.

[15] L.J. Muhammad, M.M. Islam, S.S. Usman, S.I. Ayon, Predictive Data Mining Models for Novel Coronavirus (COVID-19) Infected Patients’ Recovery, SN Comput. Sci. 1 (2020) 206. https://doi.org/10.1007/s42979-020-00216-w.

[16] T. Mahmud, M.A. Rahman, S.A. Fattah, CovXNet: A multi-dilation convolutional neural network for automatic COVID-19 and other pneumonia detection from chest X-ray images with transferable multi-receptive feature optimization, Comput. Biol. Med. 122 (2020) 103869. https://doi.org/10.1016/j.compbiomed.2020.103869.

[17] E. Luz, P.L. Silva, R. Silva, L. Silva, G. Moreira, D. Menotti, Towards an Effective and Efficient Deep Learning Model for COVID-19 Patterns Detection in X-ray Images, (2020) 1–10. https://arxiv.org/abs/2004.05717.

[18] M. Rahimzadeh, A. Attar, A New Modified Deep Convolutional Neural Network for Detecting COVID-19 from X-ray Images, (2020). http://arxiv.org/abs/2004.08052.

[19] S. Minaee, R. Kafieh, M. Sonka, S. Yazdani, G.J. Soufi, Deep-COVID: Predicting COVID-19 From Chest X-Ray Images Using Deep Transfer Learning, (2020). http://arxiv.org/abs/2004.09363.

[20] N.S. Punn, S. Agarwal, Automated diagnosis of COVID-19 with limited posteroanterior chest X-ray images using fine-tuned deep neural networks, (2020). http://arxiv.org/abs/2004.11676.

[21] A.I. Khan, J.L. Shah, M. Bhat, CoroNet: A Deep Neural Network for Detection and Diagnosis of Covid-19 from Chest X-ray Images, (2020). http://arxiv.org/abs/2004.04931.

[22] L. Wang, A. Wong, COVID-Net: A Tailored Deep Convolutional Neural Network Design for Detection of COVID-19 Cases from Chest X-Ray Images, (2020). http://arxiv.org/abs/2003.09871.

[23] Narin, C. Kaya, Z. Pamuk, Department of Biomedical Engineering, Zonguldak Bulent Ecevit University, 67100, Zonguldak, Turkey., ArXiv Prepr. ArXiv2003.10849. (2020). https://arxiv.org/abs/2003.10849.

[24] E.E.-D. Hemdan, M.A. Shouman, M.E. Karar, COVIDX-Net: A Framework of Deep Learning Classifiers to Diagnose COVID-19 in X-Ray Images, (2020). http://arxiv.org/abs/2003.11055.

[25] I.D. Apostolopoulos, T.A. Mpesiana, Covid-19: automatic detection from X-ray images utilizing transfer learning with convolutional neural networks, Phys. Eng. Sci. Med. (2020) 1–8. https://doi.org/10.1007/s13246-020-00865-4.

[26] M. J. Horry et al., “X-Ray Image based COVID-19 Detection using Pretrained Deep Learning Models,” 2020. [Online]. Available: https://engrxiv.org/wx89s/

[27] M. Loey, F. Smarandache, N.E.M. Khalifa, Within the lack of chest COVID-19 X-ray dataset: A novel detection model based on GAN and deep transfer learning, Symmetry (Basel). 12 (2020). https://doi.org/10.3390/SYM12040651.

[28] P. Kumar, S. Kumari, Detection of coronavirus Disease (COVID-19) based on Deep Features, https://Www.Preprints.Org/Manuscript/202003.0300/V1. (2020) p9. https://doi.org/10.20944/preprints202003.0300.v1.

[29] S.U.K. Bukhari, S.S.K. Bukhari, A. Syed, S.S.H. SHAH, The diagnostic evaluation of Convolutional Neural Network (CNN) for the assessment of chest X-ray of patients infected with COVID-19, MedRxiv. (2020) 2020.03.26.20044610. https://doi.org/10.1101/2020.03.26.20044610.

[30] Abbas, M.M. Abdelsamea, M.M. Gaber, Classification of COVID-19 in chest X-ray images using DeTraC deep convolutional neural network, (2020). http://arxiv.org/abs/2003.13815.

[31] Islam MZ, Islam MM, Asraf A. A combined deep CNN-LSTM network for the detection of novel coronavirus (COVID-19) using X-ray images. Inform Med Unlocked. 2020;20:100412

[32] J.P. Cohen, P. Morrison, L. Dao, COVID-19 Image Data Collection, (2020). http://arxiv.org/abs/2003.11597. (Accessed 16 July, 2020).

[33] COVID-19 chest X-ray. https://github.com/agchung (accessed July 7, 2020).

[34] Radiopaedia. COVID-19 X-ray Cases. https://radiopaedia.org/?lang=gb (Accessed 16 July, 2020).

[35] COVID-19 DATABASE |SIRM. https://www.sirm.org/en/category/articles/covid-19-database/ (accessed July 7, 2020).

[36] COVID-19 Chest X-Ray Image Repository. https://figshare.com/articles/COVID-19_Chest_XRay_Image_Repository/12580328/2 (accessed July 11, 2020).

[37] COVID-19 Image Repository. https://figshare.com/articles/COVID-19_Image_Repository/12275009/1 (accessed July 11, 2020).

[38] Mendeley Data - Augmented COVID-19 X-ray Images Dataset. https://data.mendeley.com/datasets/2fxz4px6d8/4 (accessed July 7, 2020).

[39] Chest X-Ray Images (Pneumonia) |Kaggle. https://www.kaggle.com/paultimothymooney/chest-xray-pneumonia (Accessed 16 July, 2020).

[40] NIH Chest X-rays |Kaggle. https://www.kaggle.com/nih-chest-xrays/data?select=Data_Entry_2017.csv (accessed July 7, 2020).

[41] M. Huh, P. Agrawal, A.A. Efros, What makes ImageNet good for transfer learning?, (2016). https://arxiv.org/abs/1608.08614

[42] K. Simonyan, A. Zisserman, Very deep convolutional networks for large-scale image recognition, 3rd Int. Conf. Learn. Represent. ICLR 2015 - Conf. Track Proc. (2015) 1–14.

[43] Krizhevsky, I. Sutskever, G.E. Hinton, ImageNet classification with deep convolutional neural networks, Commun. ACM. 60 (2017) 84–90. https://doi.org/10.1145/3065386.

[44] G. Huang, Z. Liu, L. Van Der Maaten, K.Q. Weinberger, Densely connected convolutional networks, Proc. - 30th IEEE Conf. Comput. Vis. Pattern Recognition, CVPR 2017. 2017-Janua (2017) 2261–2269. https://doi.org/10.1109/CVPR.2017.243.

[45] C. Szegedy, V. Vanhoucke, S. Ioffe, J. Shlens, Z. Wojna, Rethinking the Inception Architecture for Computer Vision, Proc. IEEE Comput. Soc. Conf. Comput. Vis. Pattern Recognit. 2016-Decem (2016) 2818–2826. https://doi.org/10.1109/CVPR.2016.308.

[46] U. Nazir, N. Khurshid, M.A. Bhimra, M. Taj, Tiny-Inception-ResNet-v2: Using Deep Learning for Eliminating Bonded Labors of Brick Kilns in South Asia, (2019). http://arxiv.org/abs/1907.05552.

[47] P.J. Werbos, Backpropagation Through Time: What It Does and How to Do It, Proc. IEEE. 78 (1990) 1550–1560. https://doi.org/10.1109/5.58337.

[48] Yoshua Bengio, Patrice Simard, Paolo Frasconi, Learning Long-term Dependencies with Gradient Descent is Difficult, IEEE Trans. Neural Netw. 5 (2014) 157.

[49] S. Hochreiter, The vanishing gradient problem during learning recurrent neural nets and problem solutions, Int. J. Uncertainty, Fuzziness Knowlege-Based Syst. 6 (1998) 107–116. https://doi.org/10.1142/S0218488598000094.

[50] P. Liu, X. Qiu, X. Chen, S. Wu, X. Huang, Multi-timescale long short-term memory neural network for modelling sentences and documents, Conf. Proc. - EMNLP 2015 Conf. Empir. Methods Nat. Lang. Process. (2015) 2326–2335. https://doi.org/10.18653/v1/D15-1280.

[51] M.J. Brown, L.A. Hutchinson, M.J. Rainbow, K.J. Deluzio, A.R. De Asha, A comparison of self-selected walking speeds and walking speed variability when data are collected during repeated discrete trials and during continuous walking, J. Appl. Biomech. 33 (2017) 384–387. https://doi.org/10.1123/jab.2016-0355.

[52] G.E. Hinton, N. Srivastava, A. Krizhevsky, I. Sutskever, R.R. Salakhutdinov, Improving neural networks by preventing coadaptation of feature detectors, (2012) 1–18.

[53] D.M. Hawkins, The Problem of Overfitting, J. Chem. Inf. Comput. Sci. 44 (2004) 1–12. https://doi.org/10.1021/ci0342472.

[54] D.P. Kingma, J.L. Ba, Adam: A method for stochastic optimization, 3rd Int. Conf. Learn. Represent. ICLR 2015 - Conf. Track Proc. (2015) 1–15.

[55] F. Ucar, D. Korkmaz, COVIDiagnosis-Net: Deep Bayes-SqueezeNet based diagnosis of the coronavirus disease 2019 (COVID-19) from X-ray images, Med. Hypotheses. 140 (2020). https://doi.org/10.1016/j.mehy.2020.109761.

[56] K. El Asnaoui, Y. Chawki, Using X-ray images and deep learning for automated detection of coronavirus disease, J. Biomol. Struct. Dyn. 0 (2020) 1–12. https://doi.org/10.1080/07391102.2020.1767212.

[57] X. Li, C. Li, D. Zhu, COVID-MobileXpert: On-Device COVID-19 Screening using Snapshots of Chest X-Ray, (2020). http://arxiv.org/abs/2004.03042.

[58] M.E.H. Chowdhury, T. Rahman, A. Khandakar, R. Mazhar, M.A. Kadir, Z. Bin Mahbub, K.R. Islam, M.S. Khan, A. Iqbal, N. Al-Emadi, M.B.I. Reaz, Can AI help in screening Viral and COVID-19 pneumonia?, (2020). http://arxiv.org/abs/2003.13145.

